# Impact of hospitalised patients with COVID-19 taking Renin-Angiotensin-Aldosterone System inhibitors: a systematic review and meta-analysis

**DOI:** 10.1101/2020.05.03.20089375

**Authors:** Ranu Baral, Maddie White, Vassilios S Vassiliou

## Abstract

Inhibitors of the Renin-Angiotensin-Aldosterone System (RAAS) notably Angiotensin-Converting Enzyme inhibitors (ACEi) or Angiotensin Receptor Blockers (ARB) have been scrutinised in hypertensive patients hospitalised with coronavirus disease 2019 (COVID-19) following some initial data they might adversely affect prognosis. With an increasing number of COVID-19 cases worldwide and the likelihood of a “second wave” of infection it is imperative to better understand the impact RAAS inhibitor use in antihypertensive covid positive hospitalised patients.

A systematic review and meta-analysis of ACEi or ARB in patients admitted with COVID-19 was conducted. PubMed and Embase were searched and six studies were included in the meta-analysis. Pooled analysis demonstrated that 18.3% of the patients admitted with COVID-19 were prescribed ACEi/ARBs (0.183, CI 0.129 to 0.238, p<0.001). The use of RAAS inhibitors did not show any association with ‘critical’ events (Pooled OR 0.833 CI 0.605 to 1.148, p=0.264) or death (Pooled OR 0.650, CI 0.356 to 1.187, p=0.161). In conclusion, our meta-analysis including ‘critical’ events and mortality data on patients prescribed ACEi/ARB and hospitalised with COVID-19, found no evidence to associate ACEi/ARB with death or adverse events.

## Introduction

Inhibitors of the Renin-Angiotensin-Aldosterone System (RAAS) notably Angiotensin-Converting Enzyme inhibitors (ACEi) or Angiotensin Receptor Blockers (ARB) have been scrutinised in hypertensive patients hospitalised with coronavirus disease 2019 (COVID-19) following some initial data they might adversely affect prognosis. The concern emerged due to the significant role of ACE2 as a receptor for severe acute respiratory syndrome coronavirus 2 (SARS-COV-2), enabling entry into host cells^1^. The chronic use of RAAS inhibitors, has been speculated to increase the levels of ACE2 and exaggerate the severity of COVID-19^1^. With an increasing number of COVID-19 cases worldwide and the likelihood of a “second wave” of infection it is imperative to better understand the impact RAAS inhibitor use in antihypertensive covid positive hospitalised patients. We thus conducted a systematic review and meta-analysis of RAAS blockers in patients admitted with COVID-19.

## Methods

The systematic review was conducted and reported in accordance to the Preferred Reporting Items for Systematic Reviews and Meta-Analyses (PRISMA) guidelines. Two databases, PubMed and Embase, were searched from inception to 2^nd^ May 2020 using key terms such as Angiotensin-Converting Enzyme inhibitors, Angiotensin Receptor Blockers, coronavirus disease 2019, SARS-COV-2. There were no restrictions imposed on language. A snowballing method was used to the references of retrieved papers to expand the search.

All studies identified in our search were screened using the titles and the abstracts. Duplicate studies and multiple reports from same studies were removed. Any article identified as having a potential of fulfilling our inclusion criteria underwent full-text evaluation. Any study design, except for narrative reviews or opinion-based publications, with ACEi/ARB data on adult (≥ 18 years) patients with COVID-19 was included and relevant information was extracted.

The proportion of COVID patients on ACEi/ARB and their mortality data was compared to non-ACEi/ARB patients. The data was analysed using random effects in Open Meta[Analyst] Software version 10.12 (developed by the Centre for Evidence Synthesis, Brown University, School of Public Health, Rhode Island State, USA) ^2^. Statistical heterogeneity was evaluated by calculating I^2^ statistics. The statistical significance was defined as *p*<0.05.

## Results

Our search yielded 184 studies from the database searches **(Figure 1)**. We rejected 168 trials after title-abstract screening. A total of sixteen trials underwent full-text evaluation, and seven studies ^1,3–8^ were included in meta-analysis (Table 1). Most studies were retrospective observational and multi-centre studies. Three studies that categorised ACEi and ARB use separately were included as study1 and study2.

**Figure 1:**
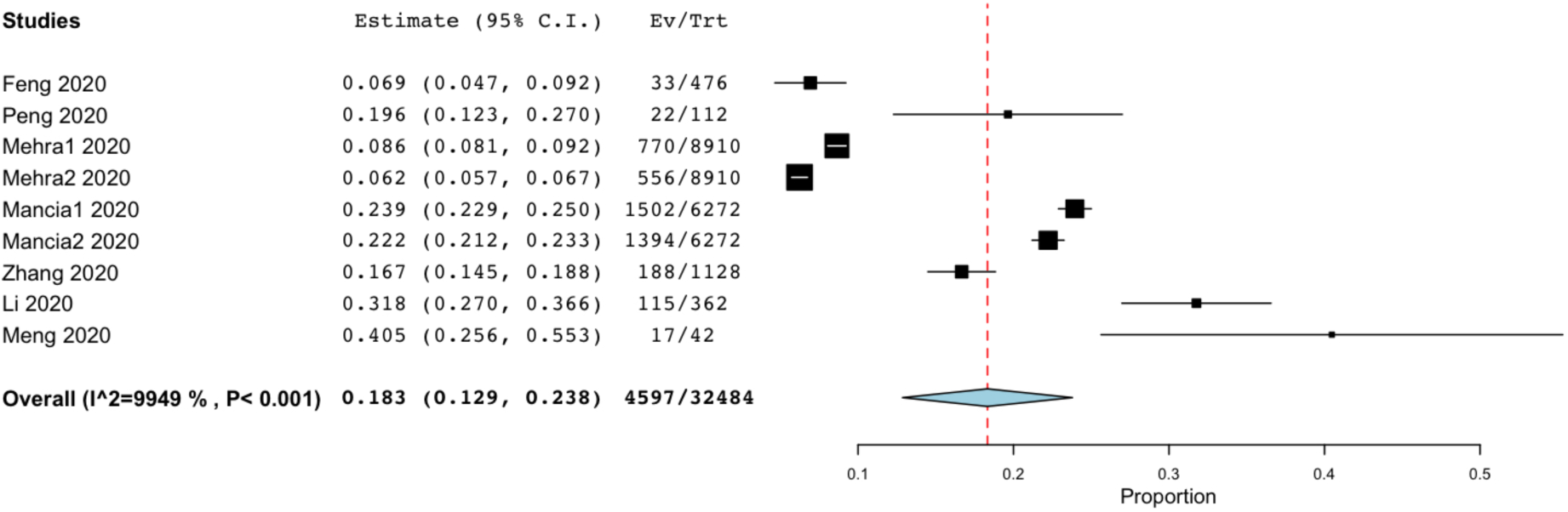
Pooled proportion of patients on ACEi/ARB with COVID-19 infection. ACEi, Angiotensin-Converting Enzyme inhibitors; ARB, Angiotensin Receptor Blockers

**Table 1:** Characteristics of included studies

| Authors | Total sample, N | ACEi/ARB, N(%) | Non-ACEi/ARB, N(%) |
| --- | --- | --- | --- |
| Li et al. | 362 | 115 (31.8) | 247 (68.2) |
| Zhang et al. | 1128 | 188 (16.7) | 940 (83.3) |
| Feng et al. | 476 | 33 (6.9) | 443 (93.0) |
| Peng et al. | 112 | 22 (19.6) | 90 (80.4) |
| Mancia et al. | 6272 | ACEi: 1502 (23.9)<br>ARB: 1394 (22.2) | Non-ACEi: 4770 (76.1)<br>Non-ARB: 4878 (77.8) |
| Mehra et al. | 8910 | ACEi: 770 (8.6)<br>ARB: 556 (6.2) | Non-ACEi: 8140 (91.4)<br>Non-ARB: 8354 (93.8) |
| Meng et al. | 42 | 17 (40.5) | 25 (59.5) |

A total of 18.3% (4597/32484) of the patients admitted with COVID-19 were prescribed ACEi/ARBs (0.183, 95% CI 0.129 to 0.238, p<0.001), as shown in Figure 1.

Some studies categorised patients as ‘critical’ or ‘severe’ which included invasive/non-invasive ventilation, intensive care unit admission or even fatalities ^6^. Reassuringly, the use of ACEi/ARB did not show any association with Covid-19 among patients who had a ‘critical’ or fatal course of the disease, Pooled OR 0.833 95% CI 0.605 to 1.148, p=0.264 as shown in Figure 2.

**Figure 2:**
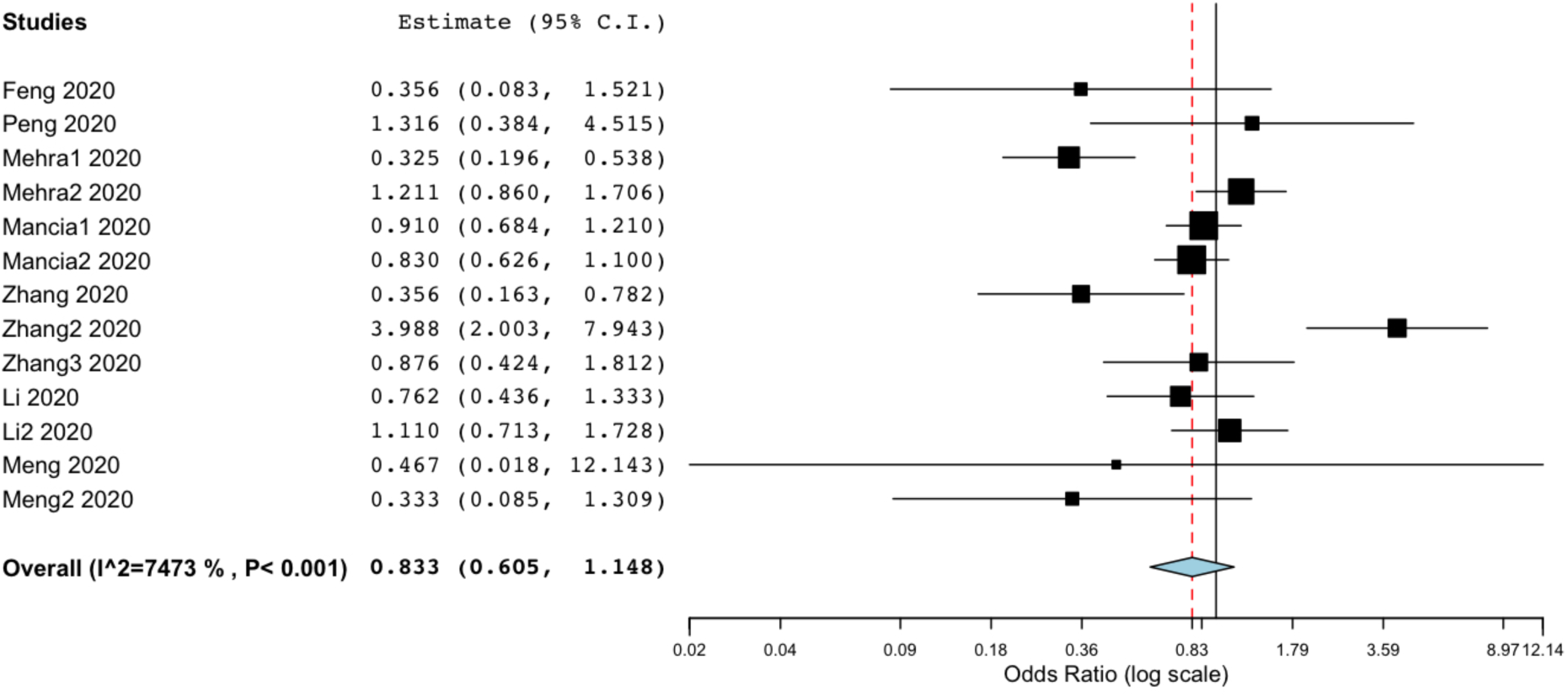
Pooled odds ratio for critical events in COVID-19 patients on ACEi/ARB including invasive/non-invasive ventilation, intensive care unit admission or death. ACEi, Angiotensin-Converting Enzyme inhibitors; ARB, Angiotensin Receptor Blockers.

Importantly, death was reported in five of the seven studies and the meta-analysis demonstrated no increased risk if taking a RAAS inhibitor; Pooled OR 0.650, 95% CI 0.356 to 1.187, p=0.161. (Figure 3).

**Figure 3:**
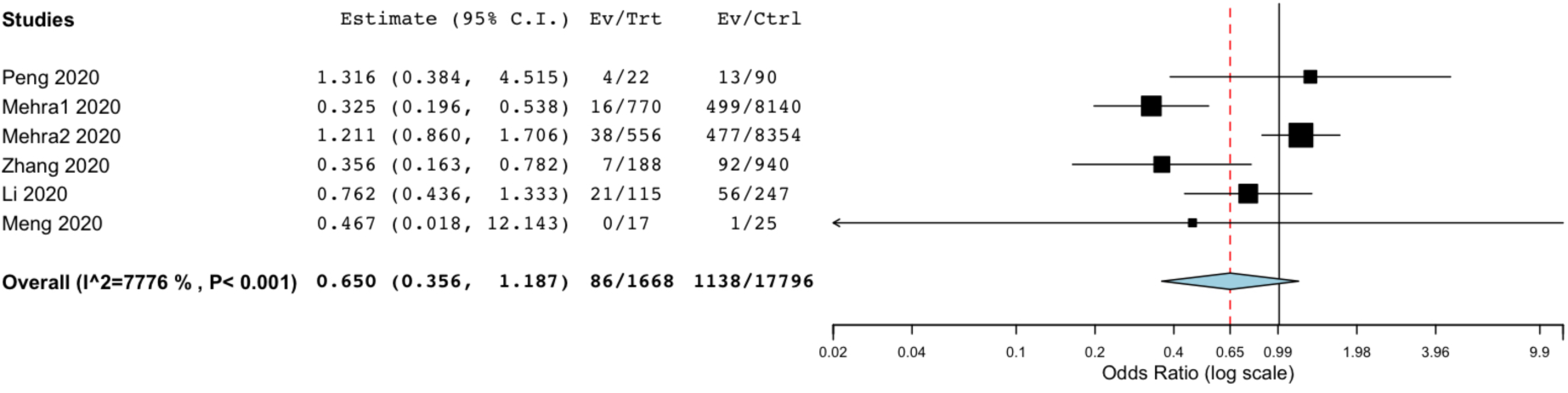
Pooled odds ratio for death in COVID-19 patients on ACEi/ARB. ACEi, Angiotensin-Converting Enzyme inhibitors; ARB, Angiotensin Receptor Blockers.

## Discussion

The uncertain role of RAAS blockers in COVID-19 has led to significant discussions and a plethora of opinions in medical communities. Whilst at present, the continued use of ACEi/ARB is advocated, there is a need for clinical data on these antihypertensives in COVID-19 patients ^9^.

Our meta-analysis showed almost a fifth of the proportion of COVID-19 patients were prescribed a RAAS inhibitor, likely due to the increasing risk of infection in patients with co-morbidities such as cardiovascular diseases, hypertension and diabetes which has been shown by previous studies^5^. Additionally, three studies ^4,5,8^ only sampled hypertensive patients with COVID.

Although cardiovascular diseases in combination with COVID-19 portend increased risk of severity and mortality^3,5^, the use of ACEi/ARB is not the likely culprit. The use of ACEi/ARB did not show any association with death among patients admitted with COVID-19. Similarly, no association was found with ‘critical’ or ‘severe’ events, including invasive/non-invasive ventilation, ICU admission or death. On the contrary, this meta-analysis showed that mortality and adverse events may even decrease with ACEi/ARB chronic use, although this did not reach statistical significance as shown in Figure 3.

Nevertheless, our study in addition to reassuring patients taking RAAS inhibitors begs an important question on whether ACEi/ARB therapy has an obscure beneficial role in patients admitted with COVID-19? Animal studies previously have shown a downregulated expression of ACE2 following SARS infection which results in increased activation of RAAS ^4,10^. This leads to a sequelae of events^4^, notably acute lung injury and consequently, adult respiratory distress syndrome (ARDS)^11^. Thus, the use of ACEi/ARB and deactivation of RAAS might be beneficial in preventing these sequence of events^4^. There are no clinical data currently on the effect of ACEi/ARB in COVID-19 and it will be interesting to see the results of further studies looking at these antihypertensives in COVID-19 patients ^12^.

### Limitations

Due to the emerging infections, there is insufficient data to compare these analyses to a control population. Heterogeneity in the meta-analysis is likely due to the varied population admitted with COVID-19 and the wide definitions of ‘critical’ or severe events.

## Conclusion

In conclusion, our meta-analysis including ‘critical events’ and mortality data on patients prescribed RAAS inhibitors and hospitalised with COVID-19, found no evidence to associate RAAS inhibition with increased deaths.

## Data Availability

All data available from the corresponding author on request

